# A comparative study of Bacterial culture isolates, site of infections and drug resistance pattern between COVID and non COVID patients admitted in a tertiary care hospital: A Pilot study

**DOI:** 10.1101/2021.09.12.21263386

**Authors:** Arup Halder, Deep Narayan Mukherjee, Soumyadeep Seal, Hindol Dasgupta, Mainak Chakraborty

## Abstract

**Introduction:** SARS-CoV2 which is a corona virus also predisposes patient to secondary bacterial infection by various mechanisms like-damaging the respiratory epithelium, profoundly affecting the innate and adaptive immunity, antagonising Interferon responses that enhance bacterial adherence, colonisation and invasion to respiratory tissue. In addition, prolonged hospital stay, invasive therapeutic devices, widespread use of empiric antibiotics and most importantly use of immune-suppressants like Steroid or Tocilizumab further increases the chances of bacterial infection. As opposed to this concept-physical distancing, frequent hand washing and use of gloves and protective gear by the healthcare workers also diminishes the chance of secondary bacterial infection. The present study is done to delineate the bacteriological profile, infection site predisposition or to gain knowledge on antibiotic sensitivity pattern.

**Method:** Retrospective data will be analyzed from June 2020, when the first COVID wave came to June 2021, corresponding to second COVID wave. The present study is a pilot study before collecting and analyzing the whole data Only those samples which were positive for bacterial isolates were randomly selected and the COVID status and drug resistance patterns were checked.

**Results and discussion:** The most common organism found was Klebsiella. Acinetobacter was also found in few patients. But most striking finding was that COVID positive patients showed higher incidence of antibiotic resistance with Acinetobacter. Though E Coli was also found commonly in COVID positive patients, they were not drug resistant.

**Conclusion:** MDR infections are common in COVID patients. Acinetobacter and Klebsiella are prone to develope MDR infections. While E.Coli is also common in COVID patients, chance of drug resistance is less among them.

## Introduction

Respiratory viral infections often predispose patients to bacterial infection [1,2]. SARS-CoV2 which is a corona virus also predisposes patient to secondary bacterial infection by various mechanisms like-damaging the respiratory epithelium, profoundly affecting the innate and adaptive immunity, antagonising Interferon responses that enhance bacterial adherence, colonisation and invasion to respiratory tissue [3,4]. In addition, prolonged hospital stay, invasive therapeutic devices, widespread use of empiric antibiotics and most importantly use of immune-suppressants like Steroid or Tocilizumab further increases the chances of bacterial infection. As opposed to this concept-physical distancing, frequent hand washing and use of gloves and protective gear by the healthcare workers also diminishes the chance of secondary bacterial infection [5].

Probably because of these reasons a controversy still exists regarding increased propensity to develop bacterial infections in COVID patients. The present study is done to fulfil the gap in the knowledge and to delineate the bacteriological profile, infection site predisposition or to gain knowledge on antibiotic sensitivity pattern. Such data can also help in the antibiotic stewardship program [6,7,8].

## Method

The study will be conducted in the department of Microbiology in a tertiary care hospital which also includes COVID care. Ethicsl committee clearance has been taken before commencing the data collection. Retrospective data will be analyzed from June 2020, when the first COVID wave came to June 2021, corresponding to second COVID wave.

The data will be analyzed statistically and any correlation or significance will be calculated with suitable statistical software. The present study is a pilot study before collecting and analyzing the whole data. The aim is to have an idea about the data trend, which may subsequently modify the data collection or interpretation methods. In the present study 30 random samples were taken from the whole period. Only those samples which were positive for bacterial isolates were selected and the COVID status and drug resistance patterns were checked. In the present study the isolates were not divided as hospital acquired or community acquired.

## Results

The total number of subjects in the present study was 30. There were 16 male and 14 female subjects. The mean age was 58.6 years (SD= 18.2767, 95% CI= 58.6 +/-6.54). among them COVID 19 Negative were 12, COVID 19 Positive were 18. The site of sample received were as follows (Table1):

**Table1.**

| Blood sample | Urine sample | Respiratory sample | Others (Ascitic) |
| --- | --- | --- | --- |
| 8 | 10 | 11 | 1 |
| 26.66% | 33.33% | 36.66% | 3.33% |

**Table 2.**

| Bacteria | Total number | COVID Positive | COVID Negative | MDR | MDR & COVID +ve | MDR & COVID -ve |
| --- | --- | --- | --- | --- | --- | --- |
| **Klebsiella** | **12 (40%)** | **5 (41.66%)** | **7 (58.33%)** | **8 (66.66%)** | **3 (37.5%)** | **5 (62.5%)** |
| E.Coli | 7 (23.33%) | 6 (85.71%) | 1 (14.28%) | 0 | 0 | 0 |
| Pseudomonus | 1 (3.33%) | 1 (100%) | 0 | 0 | 0 | 0 |
| **Acinetobacter** | **6 (20%)** | **5 (83.33%)** | **1 (16.66%)** | **3 (50%)** | **2 (66.66%)** | **1 (33.33%)** |
| Enterococcus | 1 (3.33%) | 1 (100%) | 0 | 0 | 0 | 0 |
| Proteus | 2 (6.66%) | 0 | 2 (100%) | 1 (50%) | 0 | 0 |
| Morganella | 1 (3.33%) | 0 | 1 (100%) | 0 | 0 | 0 |

The distributions of bacterial isolates among different groups were as above. Klebsiella was the most common organism recovered from the sample, while followed by E.Coli and Acinetobacter. Most of the klebsiella infections occurred among the COVID negative patients. 66.66% of Klebsiella isolates were multidrug resistant (MDR); which was again predominant in COVID negative group. 85.71% of E.Coli infection occurred in COVID positive group and none were MDR. The similar findings were present for Acinetobacter where 83.33% occurred in COVID positive group. 50% of Acinetobacter isolates were MDR and most commonly distributed in COVID positive group (66.66%).

## Discussion

The present study was done as a pilot study as described above. In this small sample size no statistical tests were performed for calculation of significance. The results are mainly tabulated as a proportion. The most common organism found was Klebsiella. Acinetobacter was also found in few patients. But most striking finding was that COVID positive patients showed higher incidence of antibiotic resistance with Acinetobacter. Though E Coli was also found commonly in COVID positive patients, they were not drug resistant.

The significance of such findings may be revealed in the bigger study planned subsequently with the same protocol. The observation of drug resistant state by each and individual bacteria in COVID positive and COVID negative patients will be an interesting finding to note; such observation may guide as to plan antibiotic therapy in COVID patients.

The scientific evidences of such data are lacking still. Previously a Spanish study on 24 subjects found most common isolates were *S. aureus, S. pneumoniae, and H*.*influenzae* [9]. Other studies also showed most common isolate was *S*.*pneumoniae* [10-13]. A furthervery small study among COVID-19 patients in ICU in Iran reported bacterial co-infection in all cases, most commonly due to Acinetobacter baumanii and possibly really representing super-infections [14]. So the organisms may vary from place to place and depend on the bacteriological profile present in a particular setup. But the data about antibiotic resistance patterns are not avidly available in COVID positive patients.

## Conclusion

MDR infections are common in COVID patients. Acinetobacter and Klebsiella are prone to develope MDR infections. While E.Coli is also common in COVID patients, chance of drug resistance is less among them.

## Supporting information

Ethics committee clearance letter

## Data Availability

Data in this study comes from a database maintained by the Brazilian Ministry of Health (MoH), which provided the anonymized database for this analyses.

## Ethics approval

This study was approved (on 01.07.2021) by Institutional

## Ethics Committee, Registration

IEC REGD NO ECR/166/Inst/Woodlands/2013/RR-20

## Notes

### Competing Interest Statement

The authors have declared no competing interest.

### Funding Statement

No funding received

### Author Declarations

Ethics approval: This study was approved (on 01.07.2021) by Institutional Ethics Committee , Registration: IEC REGD NO ECR/166/Inst/Woodlands/2013/RR-20 The approval letter is attached with the documents

## References

1. Feldman and Anderson. The role of co-infections and secondary infections in patients with COVID-19. Pneumonia (2021) 13:5 https://doi.org/10.1186/s41479-021-00083-w

2. Arnold FW, Fuqua JL. Viral respiratory infections: a cause of communityacquired pneumonia or a predisposing factor? Curr Opin Pulm Med. 2020; 26(3):208–14. https://doi.org/10.1097/MCP.0000000000000666.

3. Bengoechea JA, Bamford CGG. SARS-CoV-2, bacterial co-infections, and AMR: the deadly trio in COVID-19? EMBO Mol Med. 2020;12(7):e12560. https://doi.org/10.15252/emmm.202012560.

4. Manna S, Baindara P, Mandal SM. Molecular pathogenesis of secondary bacterial infection associated to viral infections including SARS-CoV-2. J Infect Pub

5. Murray AK. The novel coronavirus COVID-19 outbreak: global implications for antimicrobial resistance. Front Microbiol. 2020;11:1020. https://doi.org/1.3389/fmicb.2020.010200.

6. Chibabhai V, Duse AG, Perovic O, Richards GA. Collateral damage of the COVID-19 pandemic: exacerbation of antimicrobial resistance and disruptions to antimicrobial stewardship programmes? S Afr Med J. 2020; 110(7):572–3. https://doi.org/10.7196/SAMJ.2020.v110i7.14917

7. Bengoechea JA, Bamford CGG. SARS-CoV-2, bacterial co-infections, and AMR: the deadly trio in COVID-19? EMBO Mol Med. 2020;12(7):e12560. https://doi.org/10.15252/emmm.202012560

8. Hsu J. How covid-19 is accelerating the threat of antimicrobial resistance. BMJ. 2020;369:m1983.

9. Barrasa H, Martín A, Maynar J, Rello J, Fernández-Torres M, Quiñonero AA, et al. High rate of infections during ICU admission of patients with severe SARS-CoV-2 pneumonia: A matter of time? J Infect 2020. doi: https://doi.org/10.1016/j.jinf.2020.12.001.

10. Khaddour K, Sikora A, Tahir N, Nepomuceno D, Huang T. Case report: the importance of novel coronavirus disease (COVID-19) and coinfection withother respiratory pathogens in the current pandemic. Am J Trop Med Hyg. 2020;102(6):1208–9. https://doi.org/10.4269/ajtmh.20-0266.

11. Edrada EM, Lopez EB, Villarama JB, Salva Villarama EP, Dagoc BF, Smith C,Sayo AR, Verona JA, Trifalgar-Arches J, Lazaro J, Balinas EGM, Telan EFO, Roy L, Galon M, Florida CHN, Ukawa T, Villanueva AMG, Saito N, Nepomuceno JR, Ariyoshi K, Carlos C, Nicolasora AD, Solante RM. First COVID-19 infections in the Philippines: a case report. Trop Med Health. 2020;48(1):21. https://doi.org/10.1186/s41182-020-00203-0.

12. Cucchiari D, Pericás JM, Riera J, Gumucio R, Md EC. Nicolás D; hospital Clínic 4H team. Pneumococcal superinfection in COVID-19 patients: a series of 5 cases. Med Clin. 2020;155(11):502–https://doi.org/10.1016/j.medcli.2020.05.022

13. Zhu X, Ge Y, Wu T, Zhao K, Chen Y, Wu B, Zhu F, Zhu B, Cui L. Co-infection with respiratory pathogens among COVID-19 cases. Virus Res. 2020;285:198005. https://doi.org/10.1016/j.virusres.2020.198005.

14. Sharifipour E, Shams S, Esmkhani M, Khodadadi J, Fotouhi-Ardakani R, Koohpaei A, Doosti Z, EJ Golzari S. Evaluation of bacterial co-infections of the respiratory tract in COVID-19 patients admitted to ICU. BMC Infect Dis. 2020;20(1):646. https://doi.org/10.1186/s12879-020-05374-z.

