## Supplementary material for "A comparative study of Bacterial culture isolates, site of infections and drug resistance pattern between COVID and non COVID patients admitted in a tertiary care hospital: A Pilot study": Ethics committee clearance letter

INSTITUTIONAL ETHICS COMMITTEE
WOODLANDS MULTISPECIALITY HOSPITAL LTD.

WOODLANDS HOSPITAL

To

Dr Arup Halder MD (TB & RESPIRATORY MEDICINE), DCH
CONSULTANT PULMONOLOGIST
WOODLANDS HOSPITAL

**SUBJECT: IEC APPROVAL LETTER**
**(IEC REGD NO: ECR/166/Inst/Woodlands/2013/RR-20)**

A meeting of Institutional Ethics Committee of WOODLANDS MULTISPECIALITY HOSPITAL on 21.06.2021 virtually from 17.30- 19.30pm. In this meeting a research proposal title ***"A comparative study of Bacterial culture isolates, site of infection, antibiogram between COVID and non COVID patients admitted in a tertiary care hospital: Descriptive and retrospective data of a year"*** of which you are the principal investigator and Dr D.N.Mukherjee[2], Dr Soumyadeep Seal[3], Dr Hindol Dasgupta[4] Dr Mainak Chakraborty[5], Department of Clinical Microbiology, Department of Pulmonology Woodlands Multispecialty Hospital are co-investigators were reviewed.

The following additional documents submitted by you were also reviewed:

1. Study synopsis
2. Proforma for data analysis.

After deliberation and review the committee thinks the study be done in two portions; first including pilot group so as to corroborate data findings and statistical analysis more suitable. Also as retrospective patient antibiogram data belongs to hospital management, a due NOC / Allowance letter would be necessary from the Hospital management appropriate authority to initiate the study trial. NOC from the hospital management has been received and hence the committee has **APPROVED** the study to be initiated.



Scanned with CamScanner

The list of committee members present are appended herewith:

| | |
|---|---|
| Dr (Prof) S K Basu | Chairman |
| Dr. Malati Purkait | Vice Chairman |
| Dr. Soumyadeep Seal. | Member Secretary (Absent during discussion pertaining to study) |
| Dr. Anjan Adhikari | Basic medical Scientist |
| Dr. Udayan Bakshi | Clinician |
| Mr. Prabir Mitra, | Legal Expert |
| Dr. Pushpita Mandal, | Clinician |
| Mrs Sudha Kaul, | Social Scientist |
| Dr Reena Sen | Member |
| Mrs Sunita Sarkar | Lay Person |

The committee expects that any amendments to the study protocol or any other relevant documents would be brought to notice,

Date: 01.07.2021
Place: Kolkata.

Dr (Prof) S.K Basu
*Chairman*
*Institutional Ethics Committee*
*Woodlands Multispecialty Hospital*
*8/5 Alipore Road*
*Kolkata 700027*

Dr Malati Purkait
*Vice Chairman*
*Institutional Ethics Committee*
*Woodlands Multispecialty Hospital*
*8/5 Alipore Road*
*Kolkata 700027*



Scanned with CamScanner
